# An epidemiological scenario for Mass Events During the World Cup

**DOI:** 10.64898/2026.06.13.26355586

**Authors:** Jorge X. Velasco-Hernández

**Affiliations:** Instituto de Matemáticas-Juriquilla, Universidad Nacional Autónoma de México

**Keywords:** Superspreading, measles outbreak, branching process, individual reproduction number, World Cup

## Abstract

This brief work discusses potential superspreading events that may occur during the World Cup in México. The study is particularly focused on the city of Guadalajara due to a large recent outbreak in January and February and insufficient vaccine coverage previous to 2026.

## 1 Introduction

Massive events in a soccer stadium and FanFests in the streets may become superspreading events because of the high population density of these gatherings and the likely presence of individuals who are infected and, due to individual or behavioral traits, have high individual reproduction numbers. The case of interest is Guadalajara, for it is the capital of the state that was the epicenter of a large measles outbreak at the beginning of this year.

The FIFA World Cup, co-hosted by Mexico, the United States, and Canada, is being held from June 11 to July 19 of this year. During this sporting event, Mexico will host matches in three cities, for a total of 13 soccer games. The host cities will be Mexico City, Monterrey (Nuevo León), and Guadalajara (Jalisco). Due to the strong football culture, especially in Mexico and around the world, a large influx of domestic and international tourists is expected in these three cities. Table 1 taken from [1] shows the basic data of the Jalisco epidemic from January 3 to February 13, 2026.

**Table 1:**
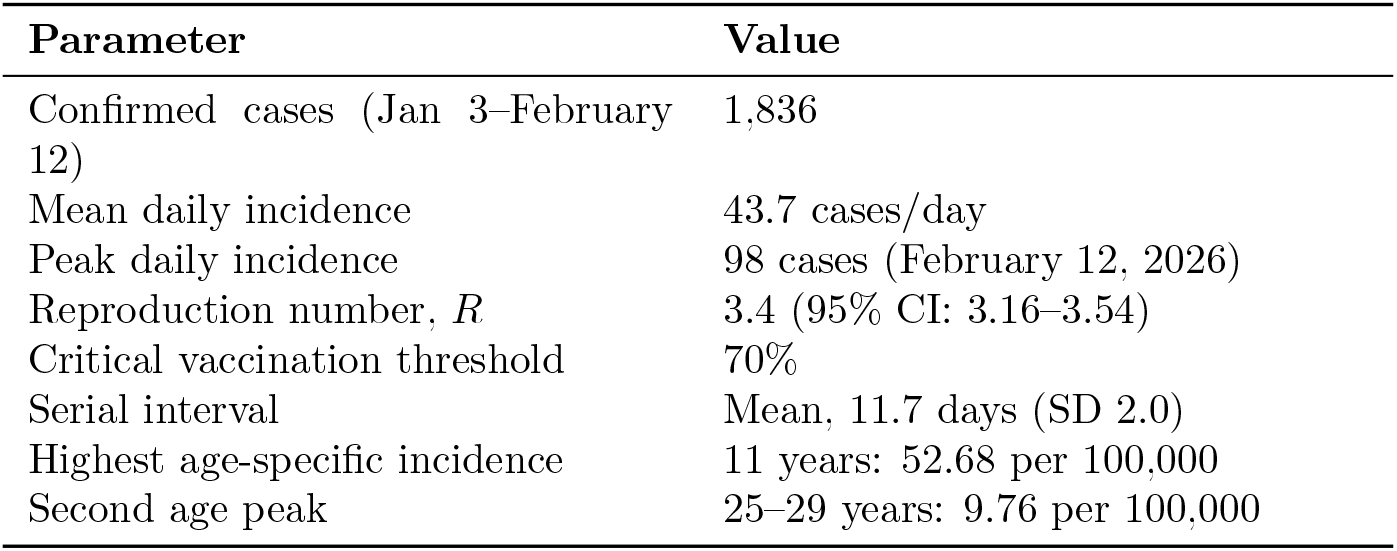
Basic indicators of measles in Jalisco 2026. Summary extracted from Subedi et al. [1].

On June 1, Lessler et al. [2] published an analysis of the likelihood of outbreaks of a set of diseases during the World Cup. In general, their prognosis regarding the importation of pathogens into the US after the World Cup is that the risk of importation to generate outbreaks is very low. However, the authors state that measles may cause outbreaks during the World Cup “[with] high consequences” (Science Adviser, June 11). These will happen in Mexico, we believe, where the risk is not that low.

## 2 Measles outbreak in Jalisco, Mexico

In this section, we briefly summarize the outbreak dynamics in the state of Jalisco, which has the highest number of cases in the country so far in 2026, and whose capital city is one of the host cities for the FIFA World Cup. We downloaded the Ministry of Health measles database, provided by the General Directorate of Epidemiology and the National Epidemiological Surveillance System (SINAVE). The following is a general summary of measles trends from January 1 to May 21, 2026. Figure 1 shows the weekly incidence and cumulative cases of measles during this period, and Table 1 reproduces the findings of a recent study on the Jalisco epidemic [1].

**Figure 1:**
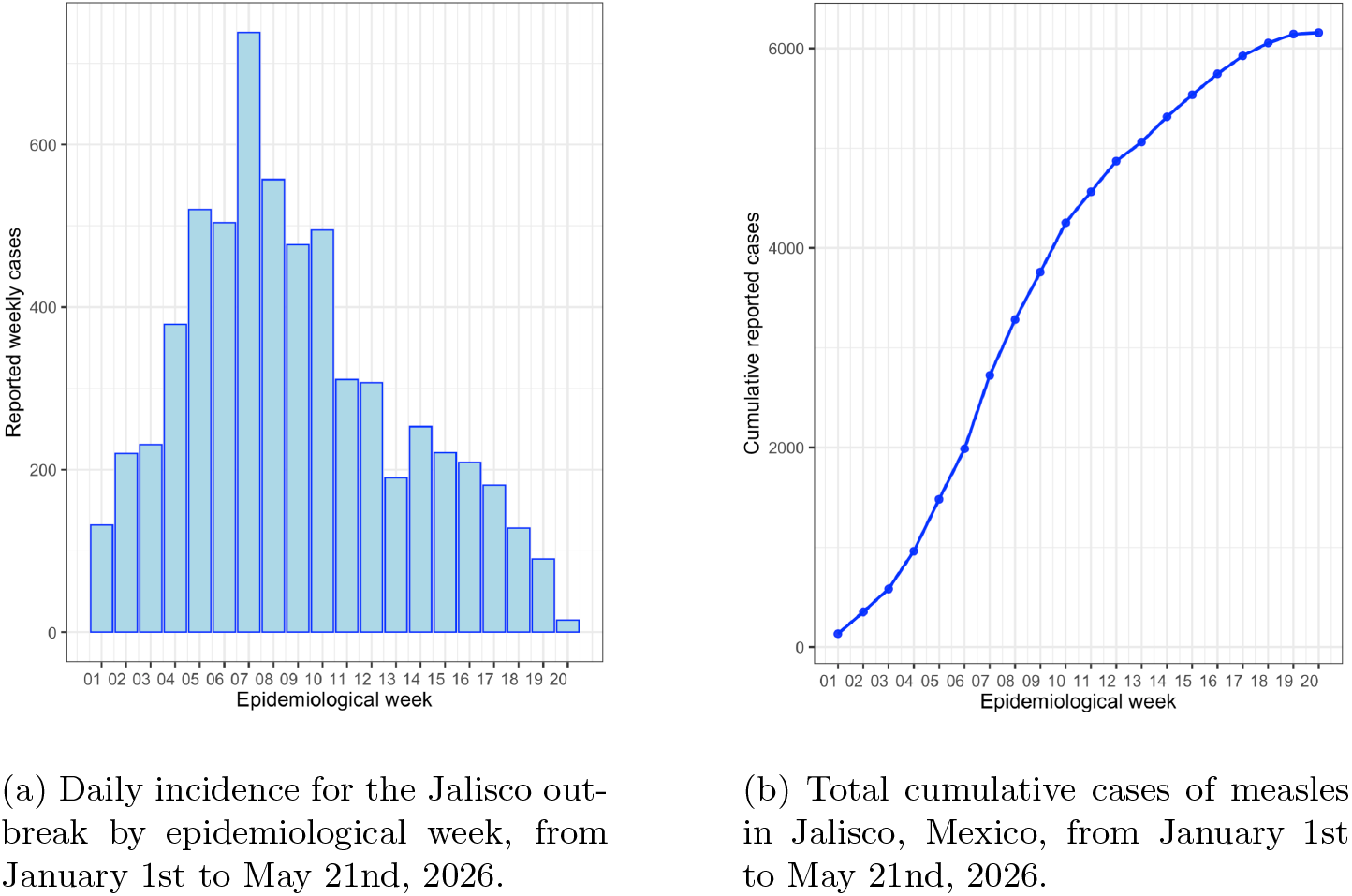
Weekly incidence and cumulative cases of measles in Jalisco, Mexico, from January 1st to May 21st, 2026.

Using [3, 4], we computed the instantaneous reproduction number shown in Figure 2.

**Figure 2:**
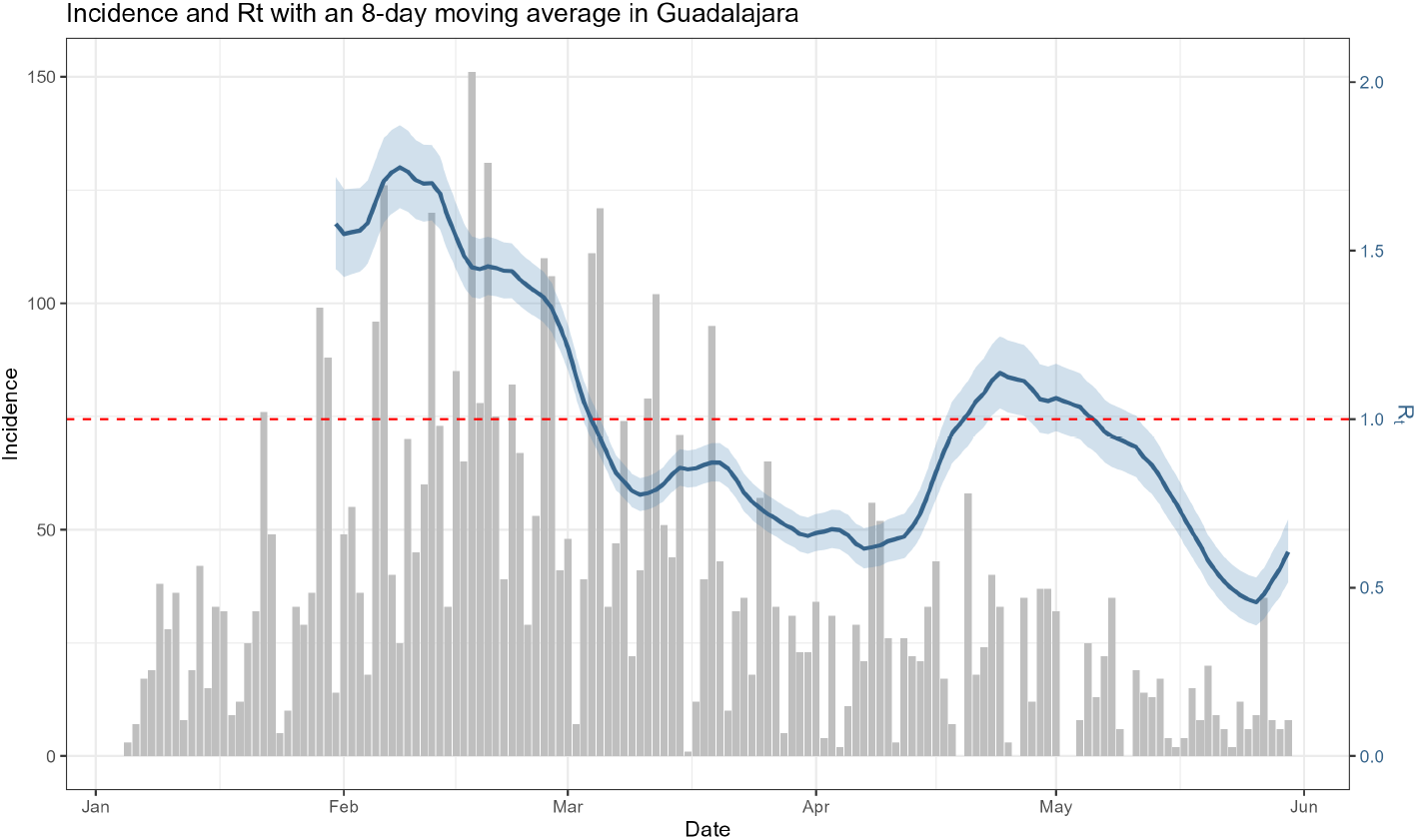
Instantaneous Reproduction Number (*R*_*t*_) for Guadalajara from February 1st to May 30. Note that we are not providing the reproductive number for the initial outbreak, as it has already been estimated in [1].

## 3 Analysis of scenarios: superspreading

Based on the information presented in the previous sections, we now present potential epidemiological scenarios arising from the arrival of domestic and international visitors for the FIFA World Cup in Mexico. The scenarios are based on the measles outbreak in the state of Jalisco, Mexico, as a reference framework.

As pointed out in [5], SARS-CoV-2 has shown that disease transmission is characterized by significant heterogeneity between individuals. Superspreader events, where some individuals infect a considerably higher proportion of the population than the average, are critical to the epidemic outcome. Superspreading is a phenomenon common to several infectious diseases, including measles. In what follows, we use the results from [6].

Accordingly, to incorporate variation in individual infectious capacities, we can assume that *ν*, the individual reproduction number, defined as the expected number of secondary cases caused by a particular infected individual [6], follows a gamma distribution with dispersion parameter *k* and mean *R*_0_, where *R*_0_ is the effective reproduction number.

### 3.1 Superspreading: scenarios for the mean effective *R*_0_

One scenario where measles outbreak risk is likely is when, due to the influx of a large number of susceptible and infected individuals from abroad (either domestic or international travelers), the effective reproduction number in Guadalajara is increased. As of 10 June, the effective reproductive number is below one, around *R*_0_ = 0.6 (see Figure 2). The following results illustrate two cases. One where the increase reaches only *R*_0_ = 1, indicating stationary transmission, and the other where there is a slight increase above threshold, with *R*_0_ = 1.5, both cases with an aggregation parameter *k* = 0.5, indicating a large variance in the distribution of individual reproduction numbers and thus high risk oLloydSmith et al [6] report that, in simulations for the latter case, 24% produce 100 first-generation infections (conditioned to non-extinction), which is a surprisingly large number given the low average reproduction number. Roughly speaking, in both scenarios (*R*_0_ = 1 and *R*_0_ = 1.5,) the individual reproduction numbers range mainly from 0 to 5 (although the dispersion is large and higher reproduction numbers are possible), with a slightly larger tail in the case of *R*_0_ = 1.5.

FIFA guidelines for seating in stadiums are as follows for general admission areas: the tread depth of the terracing should be 800 mm (minimum), with seat centers at 500 mm (minimum) in a best-practice situation. Also, in a typical arrangement of rows of seats, any person has at least eight nearest neighbors (Moore neighborhood) located between one and one and a half meters from the neighborhood center.

The Secretaría de Salud of Jalisco State reported that the state (local population)”achieved record-high measles vaccination coverage, with more than 3.3 million doses administered statewide”. Let us assume that, as of the beginning of June, coverage is around 85%-90%, which means that, for a stadium capacity of 54,000, there are between 5,400 and 8,100 unvaccinated individuals seated to watch the game. This baseline, however, is conservative because a significant proportion of the public will come from other places with lower coverage, particularly in Mexico. Assume that the percentage of unvaccinated individuals increases by around 10% to 15%, meaning there may be 10800 to 16200 nonimmune fans at the stadium (20% to 30% of the total). This is the environment in which a superdispersive event, illustrated in Figures 3 and 4, may occur.

**Figure 3:**
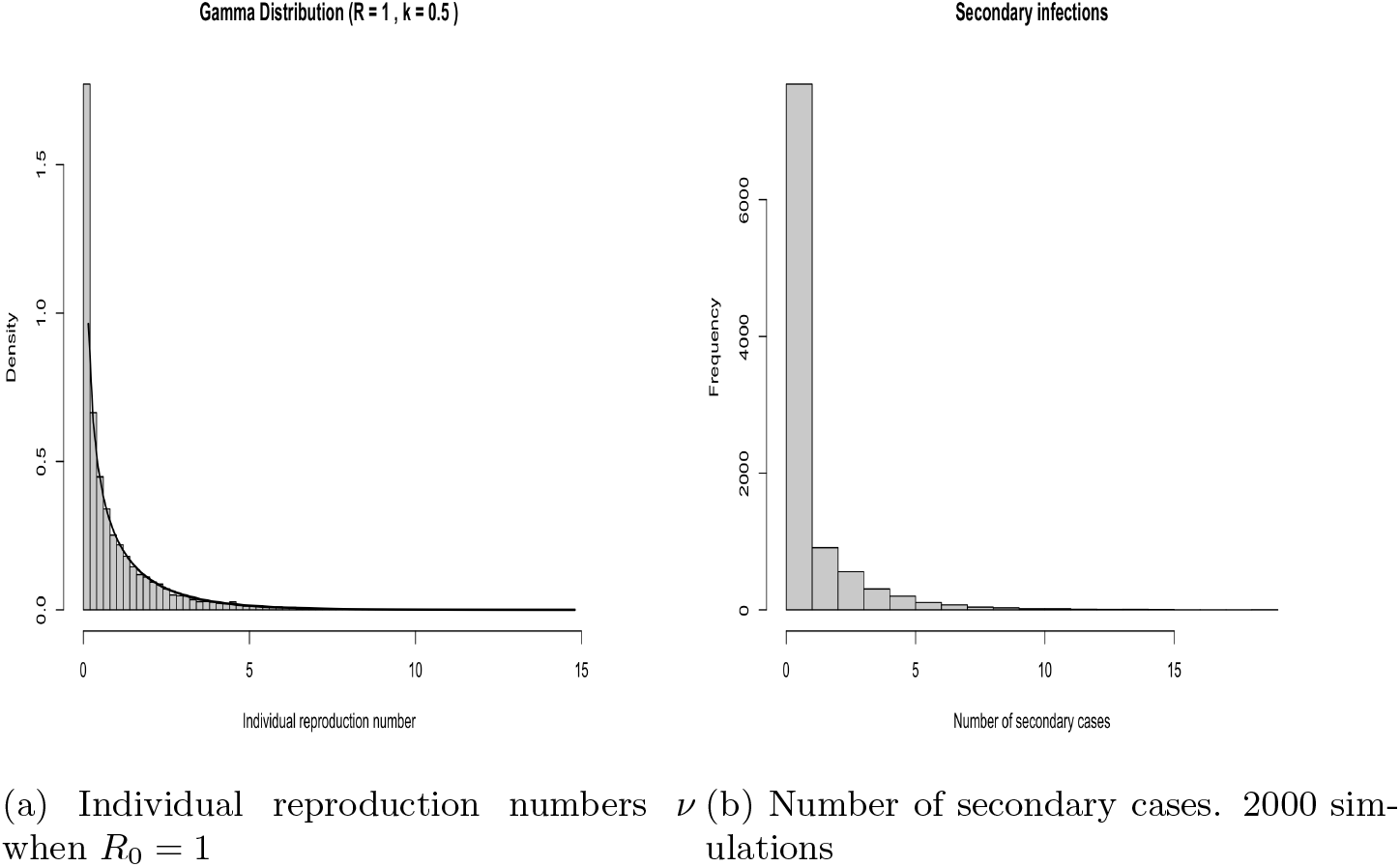
Distribution of *ν* individual reproductive numbers and the corresponding expected number of secondary cases for *R*_0_ = 1 and *k* = 0.5.

**Figure 4:**
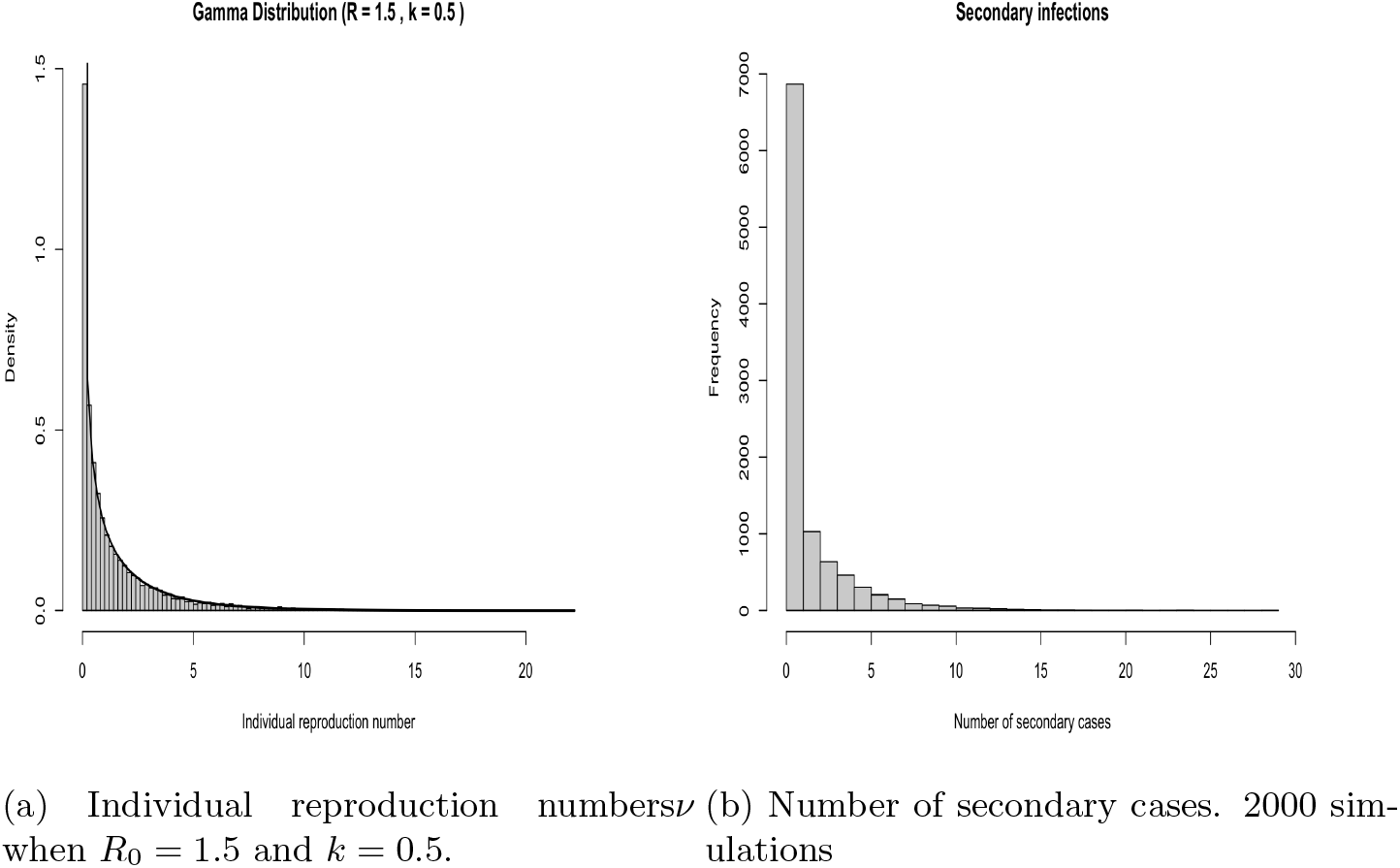
Distribution of *ν* individual reproductive numbers and the corresponding expected number of secondary cases for *R*_0_ = 1.5

### 3.2 Superspreading: how many cases?

The number of active cases in Guadalajara by the end of May is very low. The last report of active cases indicates 8 individuals. Given that under-reporting is significant and that the number of imported infectious cases is rather unknown, let us assume that 5 cases arrive at the stadium to watch the match between South Korea and Chequia. Two questions, however, still stand: Is this a realistic guess? How many non-local, actively infected individuals will arrive in Guadalajara? These are rather difficult questions to answer.

If an infected person is seated and all their contacts are vaccinated, there will be no transmission. However, any non-immune individual in that neighborhood will be infected, given measles’s high transmissibility. So, even if this infectious person is a super-spreader, the net effect will be only to infect non-immune individuals in the neighborhood. The problem arises with mobility within the stadium, particularly at the beginning, half-time, and end of the game, when people concentrate in halls and avenues to enter or leave the facilities. Here, the superspreading events, if any, could have a significant impact on prevalence later in time (after 5 to 7 days corresponding to the incubation period).

We postulate that superspreading events will undoubtedly be associated with mobility. In México, a very aggressive vaccination campaign started in 2025 but not all areas of the country have achieved the same high level of coverage as in Guadalajara.

So, using the superspreading hypothesis, we have simulated an SEIR epidemic as a branching-process to capture the nature of this phenomenon. Using the model of [6], each of the five initial cases generates secondary infections according to a Gamma-Poisson mixture distribution:

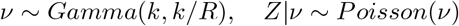

where *ν* is the individual reproduction number and *Z* the number of cases generated under the assumption of the statistical distribution of *ν*. Therefore, the offspring (secondary cases) distribution is negative binomial with mean *R*_0_ and dispersion *k* [6]. To produce the following figures, we have simulated the transmission events using a mean incubation period of 6 and a mean infection period of 5 days to approximate the reported serial interval of about 11 days for Jalisco at the beginning of this year [1] (see Table 1). We have used a 15-day time horizon after the initial match (June 11) and an initial number of 5 infected individuals. These two numbers may be varied and are reported only as examples. Given the data reported in [1], the infectious individuals are likely to be in the age brackets of 11 and [25, 29]. See Figures 5a and 5b.

**Figure 5:**
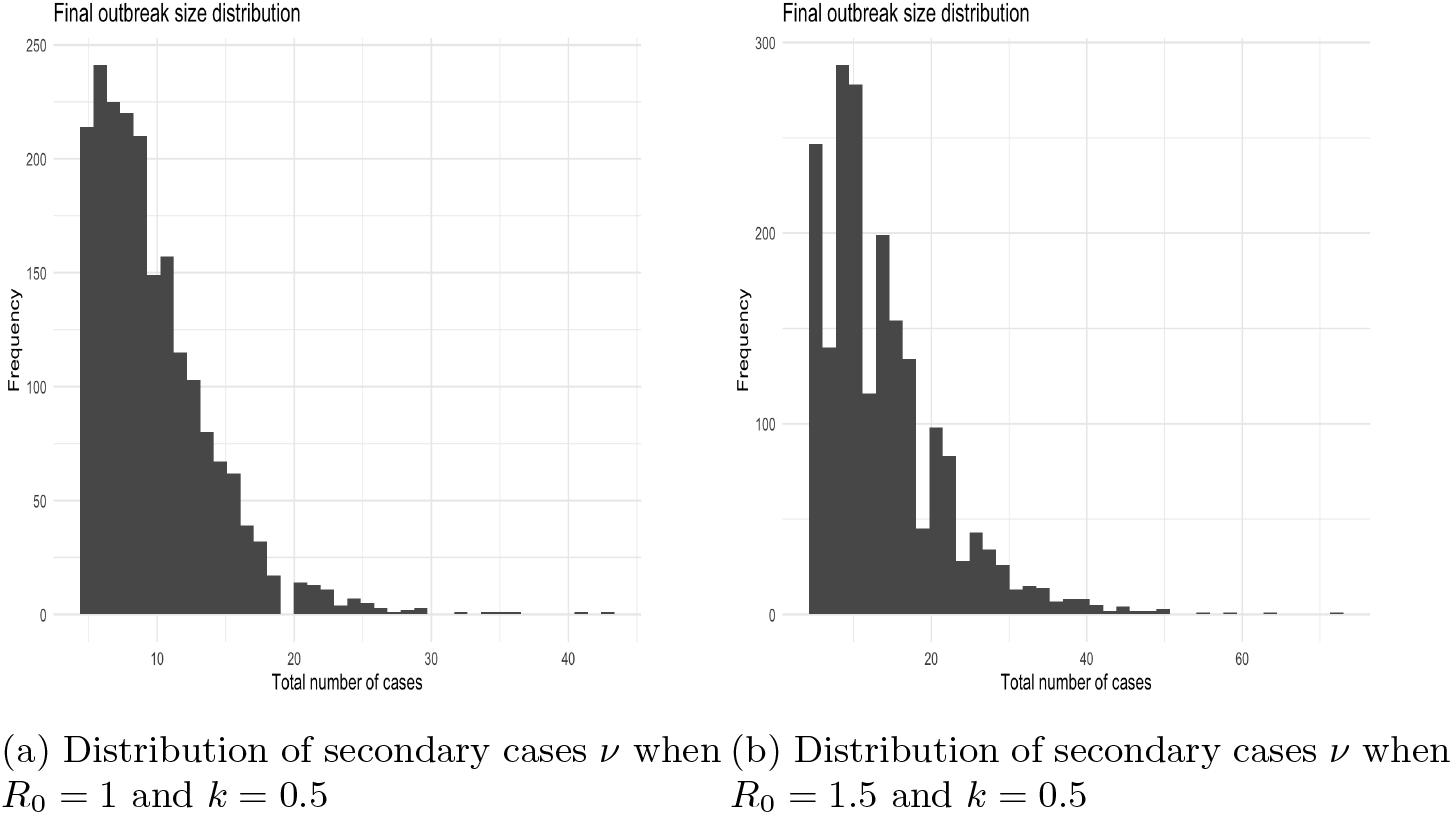
Distribution expected number of secondary cases for *R*_0_ = 1. and *R*_0_ = 1.5 with the same dispersion parameter *k* = 0.5. Number of simulations: 2000

Therefore, to reproduce the current state of the epidemic, in Figure 6 we show the same model as previously, but assuming an effective reproduction number of *R*_0_ = 0.6 and a high variance with *k* = 0.1

From these results, we can conclude that even with an average effective reproduction number below the threshold, the number of secondary cases can be rather large, with more than 30 cases for 15 days after the first match, assuming that overcrowding and mobility trigger superspreading events.

**Figure 6:**
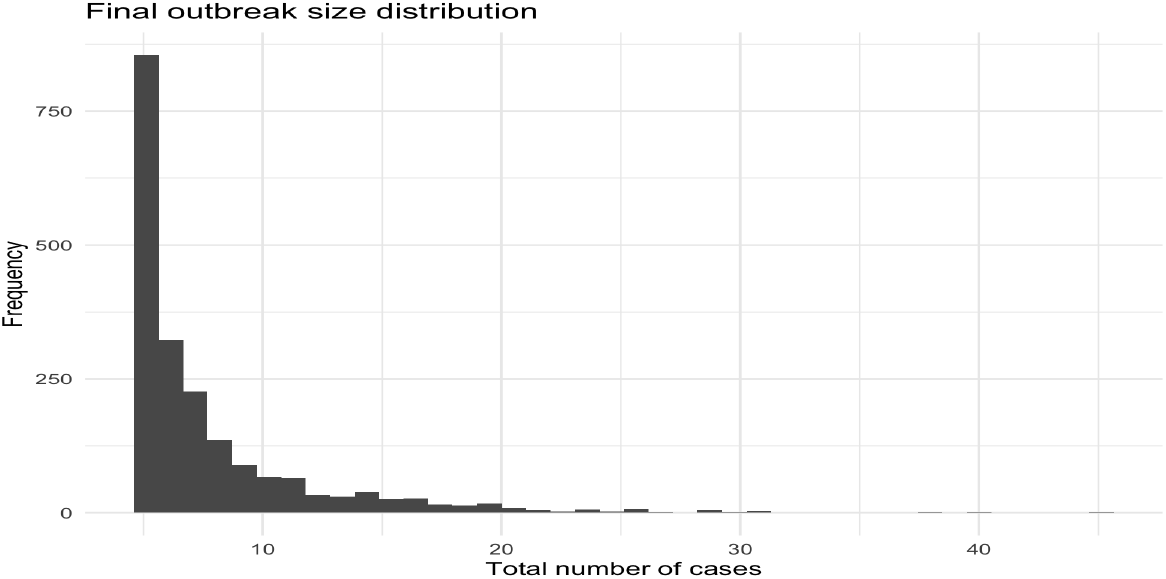
Distribution of secondary cases *ν* when *R*_0_ = 0.6 and *k* = 0.1

Finally, a useful way to communicate infection risk [7] is to quantify the probability that a gathering includes at least one infectious person, which can be interpreted as the probability of encountering an infectious individual in that crowd. Let *I*(*t*) denote the number of active infectious individuals in a locality on day *t*, and *N* (*t*) its population, which can be considered constant over time. Define

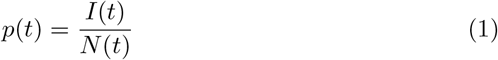

as the percentage prevalence at a site with a total of *N* individuals. Under the simplifying assumption that attendees are sampled at random and independently, the probability that a group of size *k* includes at least one infectious individual is

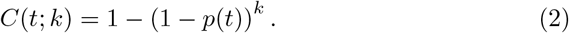

This quantity is closely related to “event risk” indicators proposed for public communication at the county level in the United States during the COVID-19 epidemic [8]. Table 2 shows the risk index for two different crowd sizes. The prevalences used are within the ranges of those reported in Mapa 1 of the “Informe Diario de Sarampión” corresponding to 8 de junio 2026 from the Dirección General de Epidemiología.

**Table 2:** Risk of contagion for a crowd size of 50000 and 30000, for different assumptions on prevalence.

| **Risk index ($k = 50000$)** | **Prevalence** |
| --- | --- |
| 0.39 | 1/100000 |
| 0.77 | 3/100000 |
| 0.91 | 5/100000 |
| 0.96 | 7/100000 |
| **Risk index ($k = 30000$)** | **Prevalence** |
| 0.25 | 1/100000 |
| 0.59 | 3/100000 |
| 0.77 | 5/100000 |
| 0.87 | 7/100000 |

## 4 Concluding remarks

This report is limited to the first match of the World Cup in Guadalajara in the state of Jalisco, which was the scenario of a large measles outbreak at the beginning of the year. One must keep in mind that in the 15 days that our projections cover, other three matches will take place in the city that have not been included in the present analysis.

The situations in Monterrey and Mexico City have distinct components that are not studied here, although superspreading events are likely to occur in both cities as well. Also, we have not dealt with the massive gatherings called Fan-Fests that will take place in the three cities. In these events, although the occupancy of the public plazas where they will take place is significantly lower than the stadium, the public is in continuous flux, and the net number of persons in attendance may be very large indeed, creating the scenario for superdispersion.

## Data Availability

All data produced are available online at
https://www.gob.mx/salud/documentos/datos-abiertos-152127

## Acknowledgments

I thank Ruth Corona Moreno for his computation of the reproduction number; I particularly thank Adrián Acuña Zegarra for useful discussions. Finally, I thank David Baca Carrasco for the initial bibliographic research on the statistics of this World Cup. This research has been developed with partial support from PAPIIT IN104726 and SECIHTI CBF-2025-G57

## References

[1] Raj Kumar Subedi et al. “A Major Epidemic of Measles in Jalisco, Mexico January - February 2026”. In: medRxiv 2026.02.17.26346510v12026.February (2026).

[2] Justin Lessler et al. Why epidemic risk at the 2026 World Cup may not be what you think. June 2026. doi: 10.64898/2026.05.28.26354384. url: http://medrxiv.org/lookup/doi/10.64898/2026.05.28.26354384.

[3] Jacco Wallinga and Marc Lipsitch. “How generation intervals shape the relationship between growth rates and reproductive numbers”. In: Proceedings of the Royal Society B: Biological Sciences 274.1609 (2007), pp. 599–604.

[4] Anne Cori et al. “A new framework and software to estimate time-varying reproduction numbers during epidemics”. In: American journal of epidemiology 178.9 (2013), pp. 1505–1512.

[5] Christopher T Lee and Thomas R. Frieden. “Identifying and Interrupting Superspreading Events—Implications for Control of Severe Acute Respiratory Syndrome Coronavirus 2”. In: Emerging infectious diseases 26 (6 June 2020), pp. 1061–1066.

[6] J. O. Lloyd-Smith et al. “Superspreading and the effect of individual variation on disease emergence”. In: Nature 438 (7066 2005). Ed. by Walter Biemel, pp. 355–359. issn: 14764687. doi: 10.1038nature04153. url: http://www.ncbi.nlm.nih.gov/pubmed/16292310.

[7] Ruth Corona-Moreno et al. “A risk-of-contagion index using a Bayesian based model for the COVID-19 epidemic in Mexico”. In: medRxiv (2026). doi: 10.64898/2026.06.09.26355274. eprint: https://www.medrxiv.org/content/early/2026/06/10/2026.06.09.26355274.full.pdf. xurl: https://www.medrxiv.org/content/early/2026/06/10/2026.06.09.26355274.

[8] Aroon Chande et al. “Real-time, interactive website for US-county-level COVID-19 event risk assessment”. In: Nature human behaviour 4.12 (2020), pp. 1313–1319.

